# Advanced Hemodynamic and Cluster Analysis for Identifying Novel RV function subphenotypes in Patients with Pulmonary Hypertension

**DOI:** 10.1101/2023.08.09.23293912

**Authors:** Alexandra M Janowski, Keeley S Ravellette, Michael Insel, Joe G Garcia, Franz P Rischard, Rebecca R Vanderpool

**Affiliations:** Division of Cardiovascular Medicine, The Ohio State University, Columbus, OH; Department of Biomedical Engineering, The Ohio State University, Columbus, OH; Division of Translational and Regenerative Medicine, The University of Arizona, Tucson, AZ; Division of Pulmonary, Allergy, Critical Care and Sleep Medicine, The University of Arizona, Tucson, AZ; Center for Inflammation Science and Systems Medicine, University of Florida

## Abstract

**Background:** Quantifying right ventricular (RV) function is important to describe the pathophysiology of in pulmonary hypertension (PH). Current phenotyping strategies in PH rely on few invasive hemodynamic parameters to quantify RV dysfunction severity. The aim of this study was to identify novel RV phenotypes using unsupervised clustering methods on advanced hemodynamic features of RV function.

**Methods:** Participants were identified from the University of Arizona Pulmonary Hypertension Registry (n=190). RV-pulmonary artery coupling (Ees/Ea), RV systolic (Ees) and diastolic function (Eed) was quantified from stored RV pressure waveforms. Consensus clustering analysis with bootstrapping was used to identify the optimal clustering method. Pearson correlation analysis was used to reduce collinearity between variables. RV cluster subphenotypes were characterized using clinical data and compared to pulmonary vascular resistance (PVR) quintiles.

**Results:** Five distinct RV clusters (C1-C5) with distinct RV subphenotypes were identified using k-medoids with a Pearson distance matrix. Clusters 1 and 2 both have low diastolic stiffness (Eed) and afterload (Ea) but RV-PA coupling (Ees/Ea) is decreased in C2. Intermediate cluster (C3) has a similar Ees/Ea as C2 but with higher PA pressure and afterload. Clusters C4 and C5 have increased Eed and Ea but C5 has a significant decrease in Ees/Ea. Cardiac output was high in C3 distinct from the other clusters. In the PVR quintiles, contractility increased and stroke volume decreased as a function of increased afterload. World Symposium PH classifications were distributed across clusters and PVR quintiles.

**Conclusions:** RV-centric phenotyping offers an opportunity for a more precise-medicine based management approach.

## Clinical Perspective

### What is new?

- Grouping participants based on pulmonary vascular resistance quintiles demonstrated an afterload-dependent increase in RV contractility and decrease in stroke volume with no change in RV-PA Coupling.
- Grouping participants using consensus clustering methods on advanced hemodynamic measures of RV function identified 5 unique clusters with distinct RV subphenotypes.
- Two RV cluster subphenotypes (C2 and C3) were identified with decreased RV contractility and RV-PA coupling when compared to participants in other clusters at a similar afterload.

### What are the clinical implications?

- Unbiased clustering approaches can help identify afterload independent RV subphenotypes that require specific therapeutic approaches.

## Introduction

Right ventricular (RV) function is a key determinant of mortality in pulmonary hypertension (PH).^1^ However, current evaluations of RV function rely on singular measures of afterload, systolic or diastolic function in addition to clinical evaluations.^2^ Maladaptation in the RV can be characterized by a combination of decreased contractility, decreased RV-pulmonary artery (PA) coupling, increased diastolic stiffness at a given RV afterload.^3^ However, a single-variable approach may lead to missing phenotypic data reflective of unique functional states. In a heterogenous disease like PH, the interplay between multiple variables can affect phenotypes in a holistic manner.^4^ When RV variables were added to current phenotyping strategies. phenotypes that reflect disease progression, patient prognosis, and highlight new potential therapeutic targets have been identified.^5,6^ Further, investigating the role that advanced RV function variables play on sub-phenotyping patients with PH may shed light on mechanisms of RV dysfunction. Functional subphenotypes that cross World Symposium Pulmonary Hypertension (WSPH) groups could give us insight into shared mechanisms complementing our current clinical classification.

Clustering analysis is an evolving tool that allows for the identification, classification, and evaluation of novel phenotypes from multiple data variables.^5,7,8^ Cluster analysis has been applied to proteomics, hemodynamics, clinical data, imaging data, and molecular and genomic data to identify novel sub-groups^7–11^ in patients with idiopathic pulmonary arterial hypertension (PAH)^5,10^, patients with exercise intolerance^7^ or patients with systemic sclerosis^11,12^.

Unsupervised clustering analysis provides an unbiased approach to holistically look at hemodynamic, RV function and clinical variables to develop phenotypes that reflect functional states of disease.^13^ Cluster analysis is fast and may identify functional signatures that may be missed by a human observer due to high-dimensional data.^14,15^ Consensus clustering evaluates multiple clustering algorithms to determine the most stable and consistent methods that best fit the dataset used.^8,16^

The aim of this study was to identify novel RV subphenotypes by applying unsupervised cluster analysis to advanced hemodynamic measures of RV function such as diastolic stiffness, contractility, and RV-PA coupling. We hypothesize that the unbiased clustering can help to provide context for advanced RV function variables in the development of RV dysfunction and the development of phenotyping strategies that are reflective of disease severity and complementary to our current forms of evaluation.

## Methods

### Patient Selection

Participants with right heart catheterization data in the University of Arizona Pulmonary Hypertension Registry were included in the analysis. Pulmonary hypertension (PH) was defined by a resting mean pulmonary arterial pressure greater than or equal to 25 mmHg in agreement with the 2015 ERS/ESC guidelines. Pulmonary arterial hypertension (PAH) was further defined by a pulmonary artery wedge pressure (PAWP) of 15 mmHg or less and pulmonary vascular resistance of 3WU or more.^17^ Participants were also classified based on WSPH classifications (WSPH group 1-5 and no PH, **Figure 1**) by an expert PH physician (FPR).^17^ Participants were excluded from analysis if they were missing RV pressure waveforms (n=23). Informed consent was obtained for the participants and was approved by the institutional review board at the University of Arizona. (IRB Protocol no. 1100000621).

**Figure. 1.**
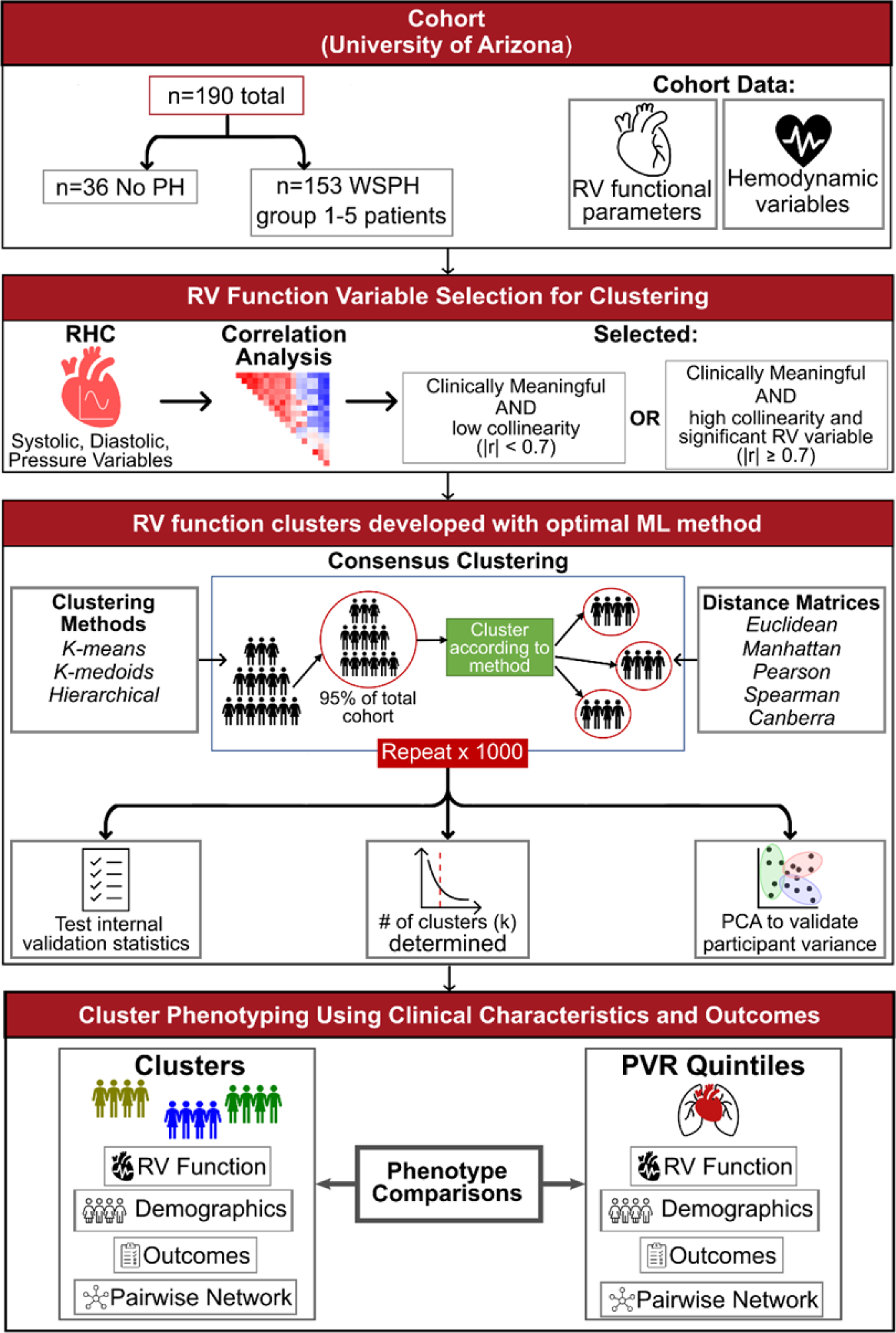
Overview of the methods used in the development of RV function clusters. Participants were identified from the University of Arizona Pulmonary Hypertension Registry (N=190). The cohort included participants with PH based on World Symposium PH guidelines (N=153, WSPH) and participants without PH (n=36, mean PA pressure <25 mmHg). RV systolic, diastolic and pressure variables were obtained from right heart catheterization data. Correlation analysis was used to reduce collinearity (|r| >0.7) and identify variables used in the clustering analysis. Clinically meaningful variables that were highly correlated with many other variables (|r|≥0.7) or only highly correlated with itself (|r|<0.7) were selected for use in clustering. Multiple unsupervised machine learning clustering methods and distance matrices were examined via consensus clustering algorithm using 95% subset of the cohort and repeated 1000 times. The optimal unsupervised machine learning method and number of clusters (k) was applied to determine distinct RV function groups based on internal validation statistics, PCA confirmation of participant variance across clusters. Clinical clustering variables were used to develop descriptions of RV function within the identified clusters and PVR quintile groups. Survival analysis was applied to determine cluster and variable associations with outcomes. RHC, right heart catheterization; PCA, principal component analysis; WSPH, World Symposium Pulmonary Hypertension groups.

### Invasive Assessment of Systolic and Diastolic Right Ventricle Function

Right heart catheterizations were performed according to standard clinical guidelines.^17^ Briefly, a pulmonary artery catheter was guided through the antecubital vein, into the right atrium, right ventricle, and pulmonary artery. Pulmonary arterial pressure (PAP), right atrial pressure (RAP), RV pressure, and pulmonary wedge pressure (PAWP) were recorded. Cardiac Output (CO) was primarily measured using thermodilution but if not available we relied on Fick cardiac output. Cardiac index was calculated as the ratio of cardiac output to body surface area (BSA). Stroke volume (SV) was calculated as cardiac output/(heart rate (HR)*1000). The pulse pressure was calculated as the difference between the systolic pulmonary arterial pressure (PAP) and diastolic PAP. Pulmonary artery (PA) compliance was calculated as the ratio of SV to pulse pressure. The contractile index was calculated as max dP/dt / systolic PAP.

Advanced measures of right ventricular function were quantified from RV pressure waveforms using the single beat method.^18–20^ Briefly, max isovolumic pressure (Pmax) was estimated from a sinusoidal curve was fit to the early isovolumic contraction and late isovolumic relaxation phase. End systolic elastance was calculated as the ratio of (max isovolumic pressure (Pmax) - RV systolic pressure (RVSP)) and SV.^21^ Arterial elastance (Ea) was calculated as the ratio between RVSP and SV. The RV-PA coupling ratio, Ees/Ea, was calculated as (Pmax-RVSP)/RVSP. RV diastolic stiffness was calculated by fitting the diastolic portion of the RV pressure-volume curve with the equation P=α(e^Vβ^-1), where α is a curve-fitting parameter and β is the diastolic stiffness coefficient.^18,19^ End diastolic elastance (Eed) is the slope at end-diastole or P = αβ(e^Vβ^). The RHC-derived stroke volume, an assumed RV end-diastolic volume of 250 ml and an RV waveform derived end diastolic pressure (RVEDP) were used to calculate Eed ^19^.

### Variable Selection and Unsupervised Consensus Clustering

Right ventricular function variables were identified for inclusion in clustering analysis. Pearson correlation analysis was used to assess collinearity. Variables with an |r|<0.7 were not considered colinear and included in the analysis. For colinear variables (|r|≥0.7), variables were grouped based on physiological meaning (systolic function, afterload, etc.) and a single representative variable was selected. Variables were then normalized to range between 0 and 1 prior to the consensus clustering.

Consensus clustering methods were used to group participants in an unbiased but rigorous approach (**Figure 1**). To identify optimal cluster stability, common unsupervised machine learning algorithms (agglomerative hierarchical, k-means, and k-medoids clustering) and distance matrices (Euclidean, Manhattan, Spearman, Pearson, Canberra) were evaluated.^22,23^ Internal validation statistics were used to determine the ideal combination of clustering method and distance matrix based on the resampling stability results (**Supplemental Methods and Table 1-3**).^24^ In the consensus clustering, each algorithm + distance combination was run on a random subsection of 95% of the cohort for each of the 1000 iterations.^25^ To determine the optimal clustering method+distance matrix combination, internal validation statistics were analyzed to find a combination that minimized the distance within clusters, the ratio of within-cluster to between cluster distance, and the widest within-cluster gap but also maximized the distance between clusters, silhouette width, separation index and the Calinski and Harabasz index stable and distinct groupings within the cohort. The optimal number of clusters (k) was evaluated for k=3-5 using the within-cluster sum of squares or “elbow” method the consensus matrix, consensus cumulative distribution functions (CDF), and cluster-consensus values (**Supplemental Figure 1**).

**Table 1:**
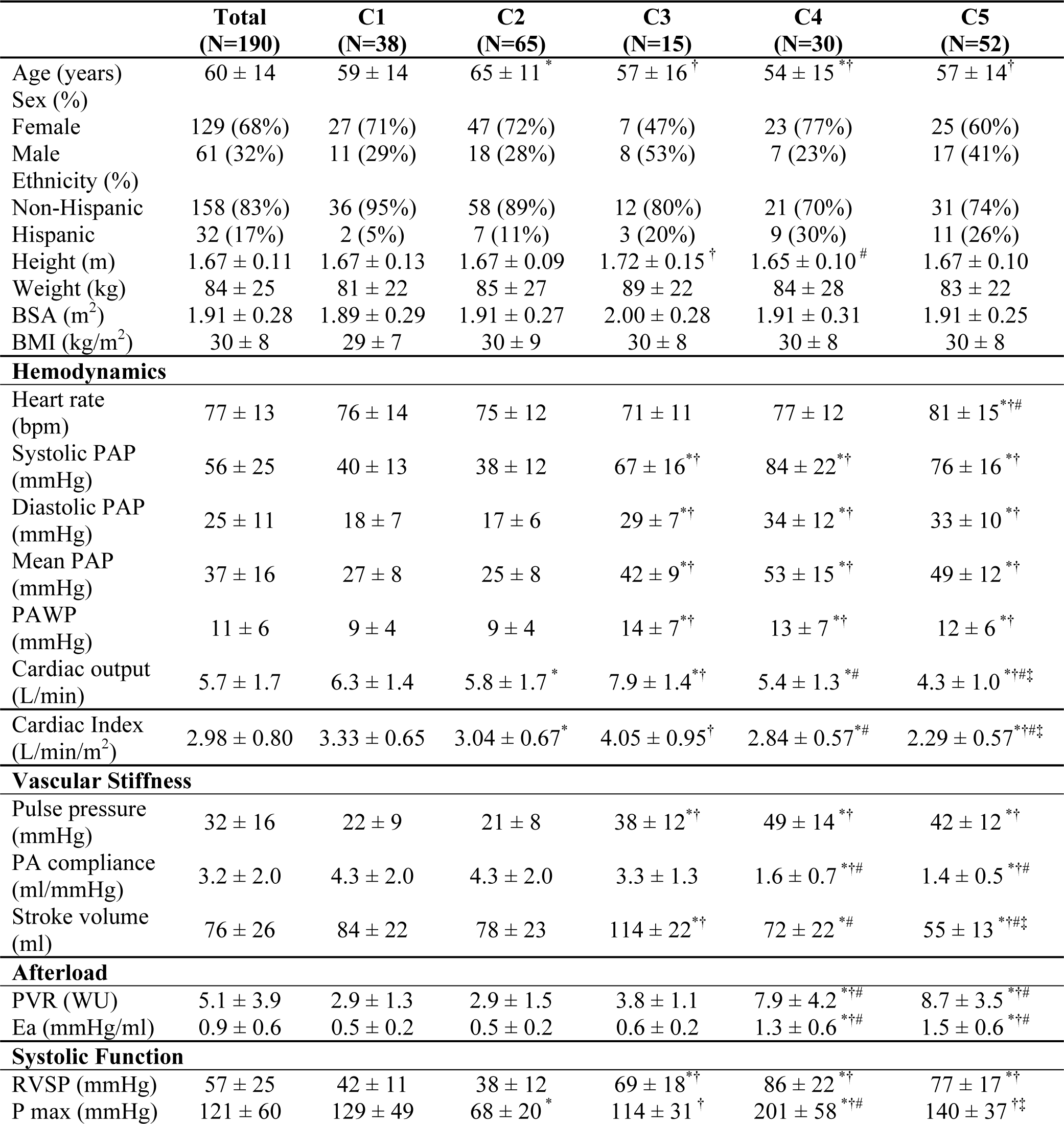
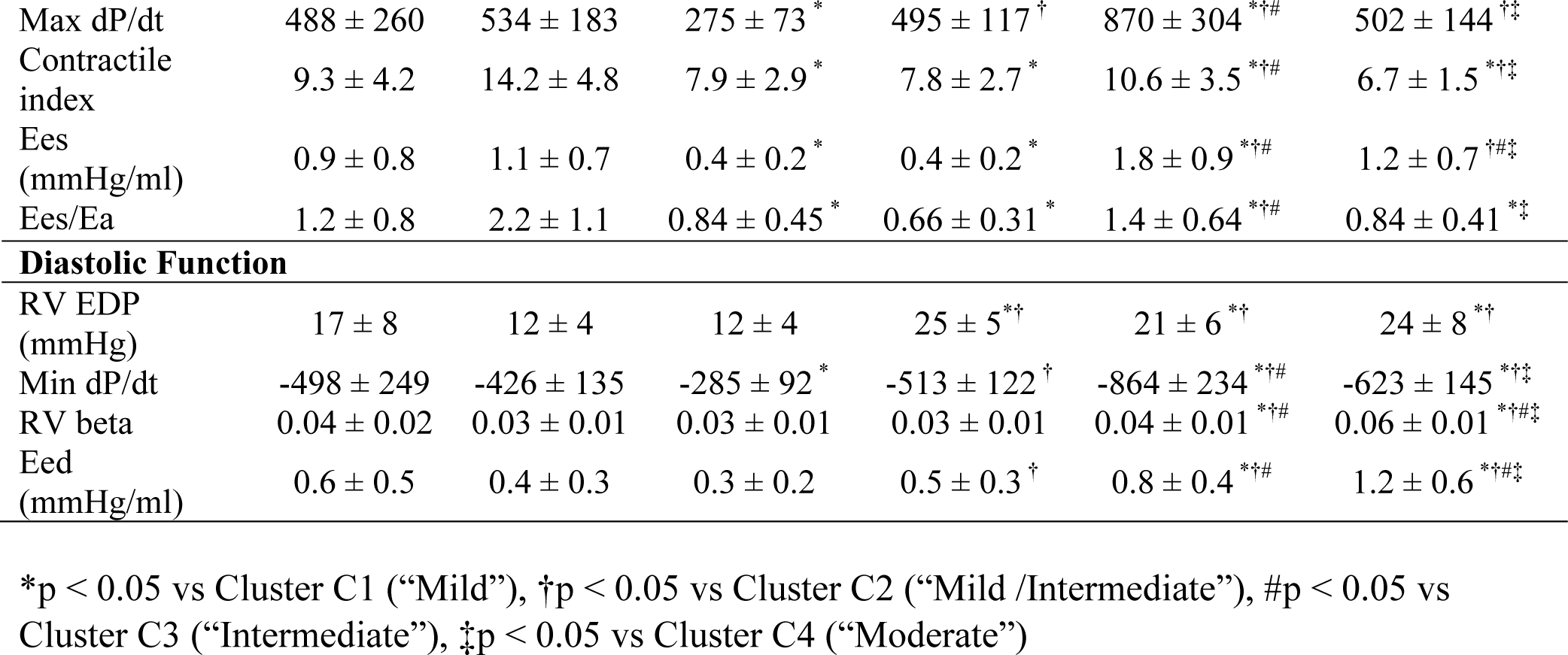
Comparison of demographics, RV function, and hemodynamics across RV function clusters. BSA, body surface area; BMI, body mass index; RVSP, right ventricular systolic pressure; RVDP, right ventricular diastolic pressure; EDP, end diastolic pressure; PAP, pulmonary artery pressure; PA, pulmonary artery; PAWP, pulmonary artery wedge pressure; BDP, beginning diastolic pressure; Eed, end diastolic elastance; Ees, end systolic elastance; Ea, arterial elastance; PVR, pulmonary vascular resistance; P max, max isovolumic RV pressure. *p < 0.05 vs Cluster C1 (“Mild”), †p < 0.05 vs Cluster C2 (“Mild /Intermediate”), #p < 0.05 vs Cluster C3 (“Intermediate”), ‡p < 0.05 vs Cluster C4 (“Moderate”)

Principal component analysis (PCA) was used to reduce the dimensionality of the variable space and visualize cluster results. Additionally, a variable correlation plot was used to investigate the relationship between variables in the first two principal components. Distance from the origin represents the quality of the variable in each principal component and the directionality of the vector is reflective of the correlation between the variable and principal component. Correlated variables are grouped together (small angle between vectors) and negatively correlated variables are in opposite quadrants (angles closer to 180 between vectors). To explore RV cluster group assignment outside of clustering, a decision tree model was constructed using the clustering variables (**Supplemental Figures 2-3**).

### Statistical Analysis

Data are presented as mean ± standard deviation for continuous variables, and as number of participants (percentage) for categorical variables. Kruskal-Wallis and Dunn tests were used to determine significance between groups for continuous variables. Chi-squared tests were used for categorical variables. A p-value of less than 0.05 was considered significant.

To visualize cluster specific phenotypic patterns, heatmap profiling and variable-variable pairwise network analysis was performed on the RV Clusters. As a comparison, the cohort was also split by PVR quintiles to visualize afterload specific heatmap and network analysis patterns. In the heatmaps, the standardized z-score for each variable was plotted for each participant and participants were grouped based on cluster assignment or separately for PVR quintiles (R-package: ‘ComplexHeatmap’)^26^. The variable-variable relationships within each cluster or PVR quintile group were investigated using pairwise network analysis. A sparse network of the top 20 partial correlations were evaluated from a weighted partial correlation network (R-packages: ‘ppcor’^27^ and ‘igraph’). Resulting networks were presented as a circle layout to highlight network variance between each cluster.

Survival analysis was performed using the time between RHC and death or follow-up (March 22, 2020). Univariable survival analysis was performed using Cox Proportional Hazards regression (p<0.1) and adjusted for age, sex, and BMI (p<0.05). Receiver Operator Characteristics (ROC) curves used to identify thresholds to be used in Kaplan Meier analyses. Differences in outcome between groups (RV function variables, clusters, and WSPH groups) were assessed using Kaplan Meier plots with pairwise Log-Rank test. The group with the greatest survival rate was assigned to be the reference group.

## Results

### Study Population

Overall, participants (n=190) were older (60±14 years), predominately female (n=129, 68%) with elevated mean pulmonary artery pressure (37±16 mmHg) and pulmonary vascular resistance (5.1±3.9 WU) (**Table 1**). Majority of the participants had PH (n=152, 81%) with elevated PVR (5.8±4 WU) compared to participants without PH (PVR: 2.0±1.0 WU, p<0.05). The majority of participants with PH had WSPH group 1 (n=92) followed by WSPH group 2 (n=22), WSPH group 3 (n=24), WSPH group 4 (n=12), WSPH group 5 (n=2) (**Supplemental Table 4**).

### Consensus Clustering Results

RV function variables that were included in the clustering analysis were Ees, Ea, Ees/Ea, Eed, Beta, RV EDP, Max dP/dt, Min dP/dt, and the contractile index. Variables with high collinearity (|r|>0.7) with other variables were excluded (RV systolic pressure, PA compliance and stroke volume). Consensus clustering internal validation statistics showed that k-medoids in combination with the Pearson distance matrix outperformed K-means and hierarchical clustering with this selection of clustering variables and cohort. (**Supplemental Table 3**). The optimal cluster number of K=5 increased cluster stability and intra-cluster consensus index values compared to K=3 or K=4 (**Supplemental Figure 1**). The five k-medoids-derived clusters were distributed across the first two principal components in distinct groups with moderate overlap (**Figure 2B**). The first and second principal components (Dim 1 and Dim 2) captured 48.7% and 23.7% of the cohort variance, respectively. All variables are contributing to cluster assignment in Dim1 and 2 (**Figure 2C**). From the variable correlation plot, diastolic variables (Ea, Eed, RVEDP, β) were grouped together with positive correlation with Dim 1 and slight negative correlation with Dim 2 (**Figure 2C**). Systolic variables congregated in two groups where max dP/dt, Ees, contractile index and Ees/Ea correlate positively with Dim 1. Min dP/dt mainly contributed to cluster assignment in Dim 1 where Max dP/dt and Ees mainly contributed to cluster assignments in Dim 2.

**Figure 2:**
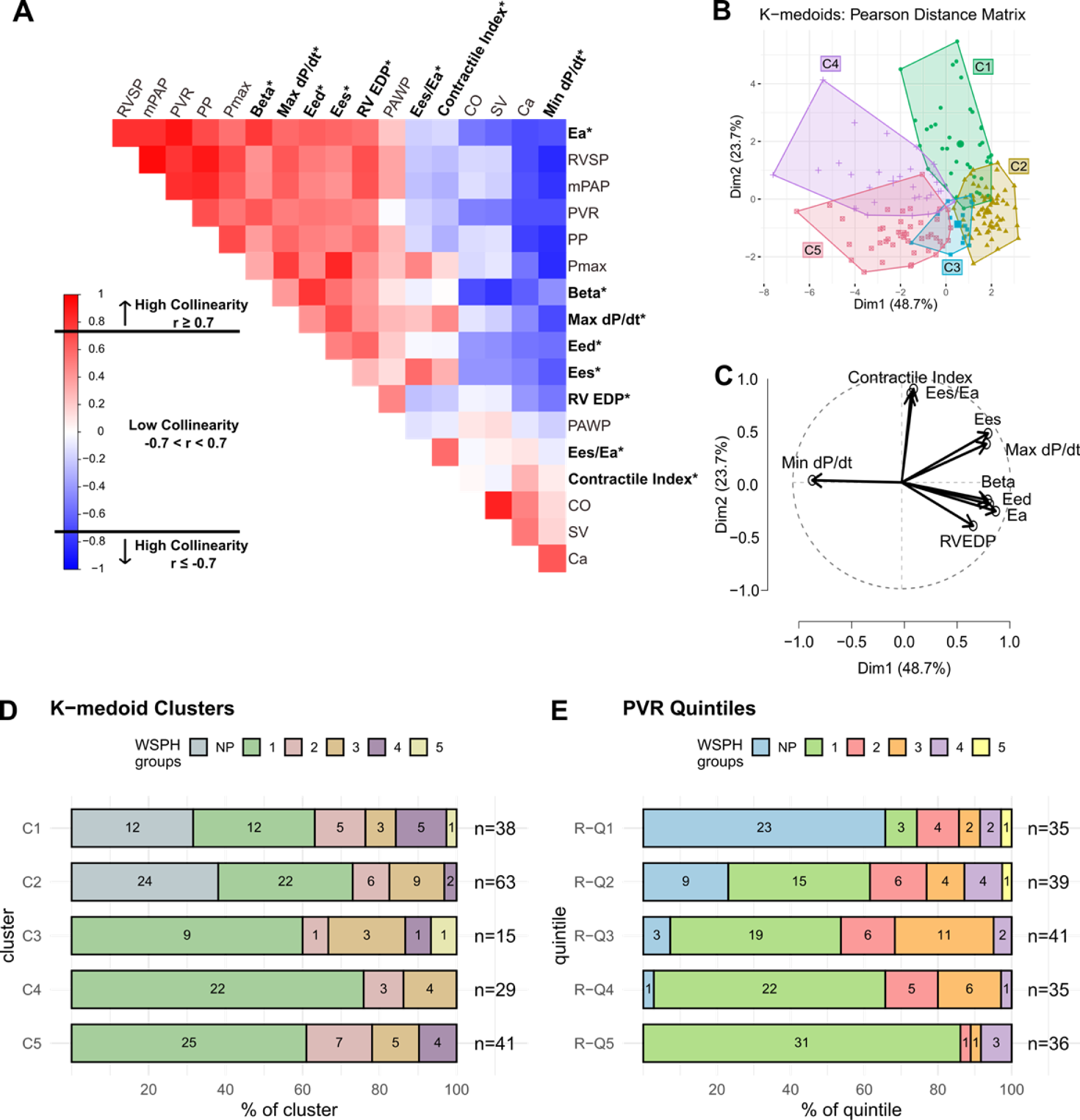
Development of RV clusters using Hemodynamic variables of RV function. **A)** Pearson correlation analysis identified variables with low collinearity (|r| < 0.7) including: Ees/Ea and the contractile index(red: positive correlations and blue: negative correlations). Variables with high correlations (|r| > 0.7) but are clinically meaningful were also considered in the clustering analysis. The chosen variables are designated an * by their labels. **B)** K-medoid cluster assignment (C1, C2, C3, C4 and C5) projected on the first two principal component analysis plot. The first two principal components (Dim 1 and Dim 2) account for 72.4% of the variance. Clusters are in distinct groups with some overlap between C2, C3, and C5. Each symbol represents a single participant. Colors and areas represent cluster assignment. **C)** Variable Correlation plot shows that all variables are contributing to cluster assignments in Dim 1 and 2. Diastolic function variables and the systolic function variables are grouped respectively. The majority of Min dP/dt contribution to cluster assignment is in Dim 1 where Contractile index and Ees/Ea mainly contributed in Dim 2. **(D and E)** World Symposium Pulmonary Hypertension (WSPH) groups distributed across RV function cluster (**D**) or PVR quintile (**E**). Each bar shows number of participants from each WSPH group in each cluster (C1 to C5) and PVR quintile (R-Q1 to R-Q5). is representative of the percentage of participants in each WSPH group. WSPH groups are well distributed across both clusters and PVR quintiles, suggesting that WSPH does not reflect cluster assignment or degree of afterload on the right ventricle. **Abbreviations:** Ea, arterial elastance; PVR, pulmonary vascular resistance; mPAP, mean pulmonary arterial pressure; PP, pulse pressure; Pmax, max isovolumetric RV pressure; Ees, end systolic elastance; Eed, end diastolic elastance; EDP, end diastolic pressure; PCWP, pulmonary capillary wedge pressure; BDP, beginning diastolic pressure; Ees/Ea, RV-PA coupling ratio; CO, cardiac output; SV, stroke volume; Ca, pulmonary artery compliance.

### Characterizing RV function in identified Clusters

Sex, BSA, and PAWP are heterogenous throughout all the clusters (**Table 1**). There were no significant differences in age between clusters except participants in Cluster C2 are significantly older than the other clusters (age = 65±11 years, p=0.001). Analysis of the clustering variables across clusters suggest the clusters can be grouped into RV subphenotypes with characteristics of reduced RV function including decreased contractility and RV-PA coupling (**Figure 3 and Table 1**).

- **Cluster 1** (C1, n = 38): Pulmonary pressure (p<0.05 vs C3-C5) and afterload (PVR 2.9±1.3 WU, p<0.05 vs C4-C5) are low compared to other clusters. Additionally, RV diastolic stiffness is low (Eed: 0.4±0.3 mmHg/ml, p<0.05 vs C4-C5) and systolic function is high (Ees/Ea: 2.2±1.1, p<0.05 vs C4-C5). Cardiac output (CO: 6.3±1.4 L/min, p<0.05 vs C4-C5) and PA compliance (Ca: 4.3±2.0 ml/mmHg, p<0.05 vs C2-C5) are good.
- **Cluster 2** (C2, n = 65): Characterized by decreased RV contractility (Ees: 0.4 ± 0.2 mmHg/ml, p < 0.05 vs C1, C4-C5) and RV-PA coupling ratio (Ees/Ea: 0.84±0.45, p < 0.05 vs C1 and C4) despite low pulmonary artery pressure (p<0.05 vs C3-C5), and low afterload (PVR 2.9±1.5 WU, p<0.05 vs C4-C5). RV diastolic function (Eed: 0.4±0.3 mmHg/ml, p<0.05 vs C3-C5). Identifying participants with decreased RV function relative to the pulmonary afterload.
- **Cluster 3** (C3, n = 15): Pulmonary pressure is increased (mPAP: 42±9 mmHg, p<0.05 vs C1-2) but with a similar RV function profiles as Cluster 2. RV function is decreased with decreased RV contractility (p<0.05 vs C1, C4-C5) and RV-PA coupling ratio (p<0.05 vs C1 and C4). Stroke volume (SV: 114±22ml, p<0.05 vs all other groups) and cardiac output (CO: 7.9±1.4 L/min, p<0.05 vs all other groups) are the highest relative to other clusters.
- **Cluster 4** (C4, n = 30): In combination with increased afterload (Ea: 1.3±0.6 mmHg/ml, p<0.05 vs C1-C3), RV contractility (Ees: 1.8±0.9 mmHg/ml, p<0.05 vs all other groups) is increased compared to the other clusters. RV diastolic stiffness (Eed: 0.8±0.4, p<0.05 vs C1-C3) is also increased. These measures of RV function are suggestive of an adapted RV with preserved RV-PA coupling (Ees/Ea: 1.4±0.64, p<0.05 vs all other groups)..
- **Cluster 5** (C5, n = 42): Decreased RV contractility (Ees: 1.2±0.7 mmHg/ml, p<0.05 vs C2-C4) and decreased RV-PA coupling (Ees/Ea: 0.84±0.41, p<0.05 vs C1 and C4) compared to Cluster 4 suggest decreased RV function. Further elevation in RV diastolic stiffness (Eed: 1.2±0.6 mmHg/ml, p<0.05 vs all other groups) is potentially suggestive of more RV failure.

**Figure 3:**
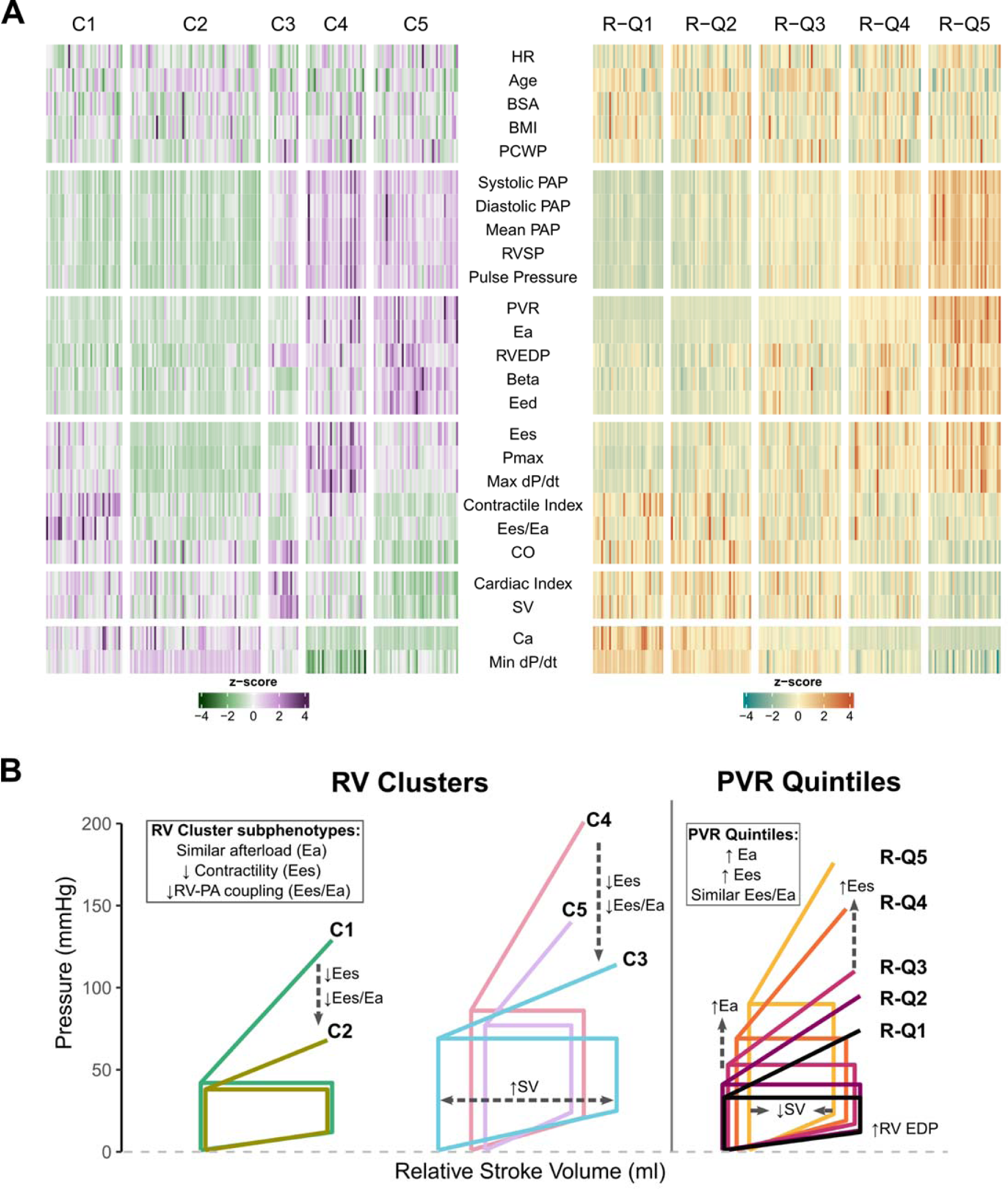
Representation of RV function profiles determined through clustering and by pulmonary vascular resistance. **(A)** Heatmap of RV function, demographic, and hemodynamic variables (rows) in relation to individual participants grouped by k-medoids cluster (C1-C5) or PVR quintile (R-Q1 – R-Q5). Z-score represents each value as whether it is greater than or less than the overall cohort average. Each cluster is reflective of a distinct functional phenotype. The clusters have more nuance in function than the PVR quintiles, which primarily reflects functional extremes. C3 uniquely has high CO that is not reflected in the other clusters/quintiles. (**B**) Representative pressure-volume loops for each cluster (left) and PVR quintile (right) based on the mean values for stroke volume, RV end-diastolic pressure, RV systolic pressure and Pmax. The pressure-volume loops in the RV clusters demonstrate the clusters 2, 5 and 3 have decreased contractility and RV-PA coupling (Ees/Ea) compared to cluster C1 and C4. Cluster 3 has an increase in stroke volume compared to all other clusters. Pressure-volume loop analysis for the PVR quintiles demonstrate that RV contractility (Ees) and RV end-diastolic pressure increase with increased afterload (Ea). There was a decrease in stroke volume going from R-Q1 to R-Q5. There were no significant changes in RV-PA coupling (Ees/Ea) in the PVR quintiles. Abbreviations: BSA, body surface area; BMI, body mass index; RVDP, RV diastolic pressure; HR, heart rate; RVSP, RV systolic pressure; RVEDP, RV end diastolic pressure; PAP, pulmonary artery pressure; PVR, pulmonary vascular resistance; Pmax, max isovolumic RV pressure; Ea, arterial elastance; Eed, end diastolic elastance; Ees, end systolic elastance; Ees/Ea, RV-PA coupling ratio; PAWP, pulmonary artery wedge pressure; CO, cardiac output; SV, stroke volume; Ca, PA compliance

Representative pressure-volume loops for the RV clusters (**Figure 3B**) demonstrate that Cluster 2 and cluster 3 have decreased contractility and RV-PA coupling compared to the other clusters. The increased stroke volume despite decreased Ees and Ees/Ea is suggestive of a shift to the Frank Starling mechanism for maintaining cardiac output in cluster 3.

### RV function subphenotypes in Pulmonary Vascular Resistance groups

As a comparison, the cohort was split into PVR quintiles with cut points of 2.1WU, 3.2 WU, 4.7 WU, and 8.0 WU (**Figure 3A, Supplemental Table 5**). RV function in the PVR groups (R-Q1 to R-Q5) is reflective of the increased afterload (**Figure 3A, right**). In R-Q1 (PVR: 1.5±0.4, n = 35), pulmonary pressure (mPAP: p<0.05 vs all other quintiles) and diastolic function (Eed: 1.1±0.5 mmHg/ml, p<0.05 vs R-Q3, R-Q4, and R-Q5) are low. There were no significant differences in Ees/Ea between quintiles (Kruskal Wallis p = 0.43, **Supplemental Table 5**). In R-Q2 (PVR: 2.5±0.3 WU, n = 40), contractile index (p<0.05) and min dP/dt (p<0.05) are decreased with no other significant changes in RV function variables compared to R-Q1. Signs of RV dysfunction start to emerge in R-Q3 (PVR: 4.0±0.5, n = 41) with increased Eed (0.6±0.4, p<0.05 vs R-Q1 and R-Q2) and decreased contractile index (8.7±3.4 p<0.05 vs R-Q1). RV diastolic dysfunction is more pronounced in R-Q4 and R-Q5 quintiles with a further increase in Eed. As expected, PA compliance and stroke volume decreased in response to increased PVR. Ees increased for both R-Q4 and R-Q5 but with no change in Ees/Ea suggesting preserved RV-PA coupling. This is in contrast to the cluster groups that have more distinct RV function phenotypes and RV-PA coupling (**Figure 3A, left and Figure 3B, left**). Representative pressure-volume loops for the PVR quintiles (**Figure 3B**) demonstrate increased RV contractility (Ees), increased RV end-diastolic pressure and decreased stroke volume with the increase in afterload from R-Q1 to R-Q5. No significant changes in Ees/Ea between PVR quintiles.

### RV Clusters and WSPH classifications

Grouping participants based on WSPH classifications does not lend itself to great phenotypic assessment of RV function (**Supplemental Table 4**). Participants with WSPH group 1 have increased PVR compared to WSPH groups 2-4 with similar measures of RV function between WSPH groups (**Supplemental Table 4**). WSPH group 3 had decreased RVSP, Pmax, and Ees compared to WSPH group 1 but no significant differences in Ees/Ea. There were no significant differences in RV diastolic stiffness between WSPH groups.

WSPH groups were distributed across the K-medoid clusters and PVR quintiles (**Figure 2D and E**). There were significantly more participants with no PH in Clusters C1 and C2 (p = 0.0005). WSPH groups 1-4 were distributed across all clusters with the largest proportion of participants with WSPH group 1 in clusters C3–C4. When split by PVR quintiles, participants without PH were primarily in R-Q1 with decreasing numbers in other quintiles. Whereas, the percentage of participants with WSPH group 1 increases from R-Q1 to R-Q5. WSPH groups 2-4 are distributed across the PVR quintiles.

### Variable-Variable Interactions within Clusters and Quintiles

Variable-variable networks were developed to investigate the variable cross-talk in the clusters and PVR quintiles (**Figure 4**). In cluster 1, the prominent interactions were observed between hemodynamic and RV systolic variables, while interactions with RV diastolic variables were limited (**Figure 4, C1**). In contrast, cluster 2 (C2) had increased cross-talk between RV systolic and hemodynamic variables. For cluster 3 (C3), there were strong interactions between all hemodynamic, RV systolic and RV diastolic variables. Variable interactions in cluster 4 (C4) have a similar pattern as C2 but with more interactions within RV diastolic variables. In cluster 5 (C5), there was an increased degree of cross-talk between hemodynamic and RV diastolic function variables but with minimal cross-talk hemodynamic and RV systolic function variables. Variable-variable interactions within the clusters do not necessarily suggest a continuum of afterload-induced RV dysfunction.

**Figure 4:**
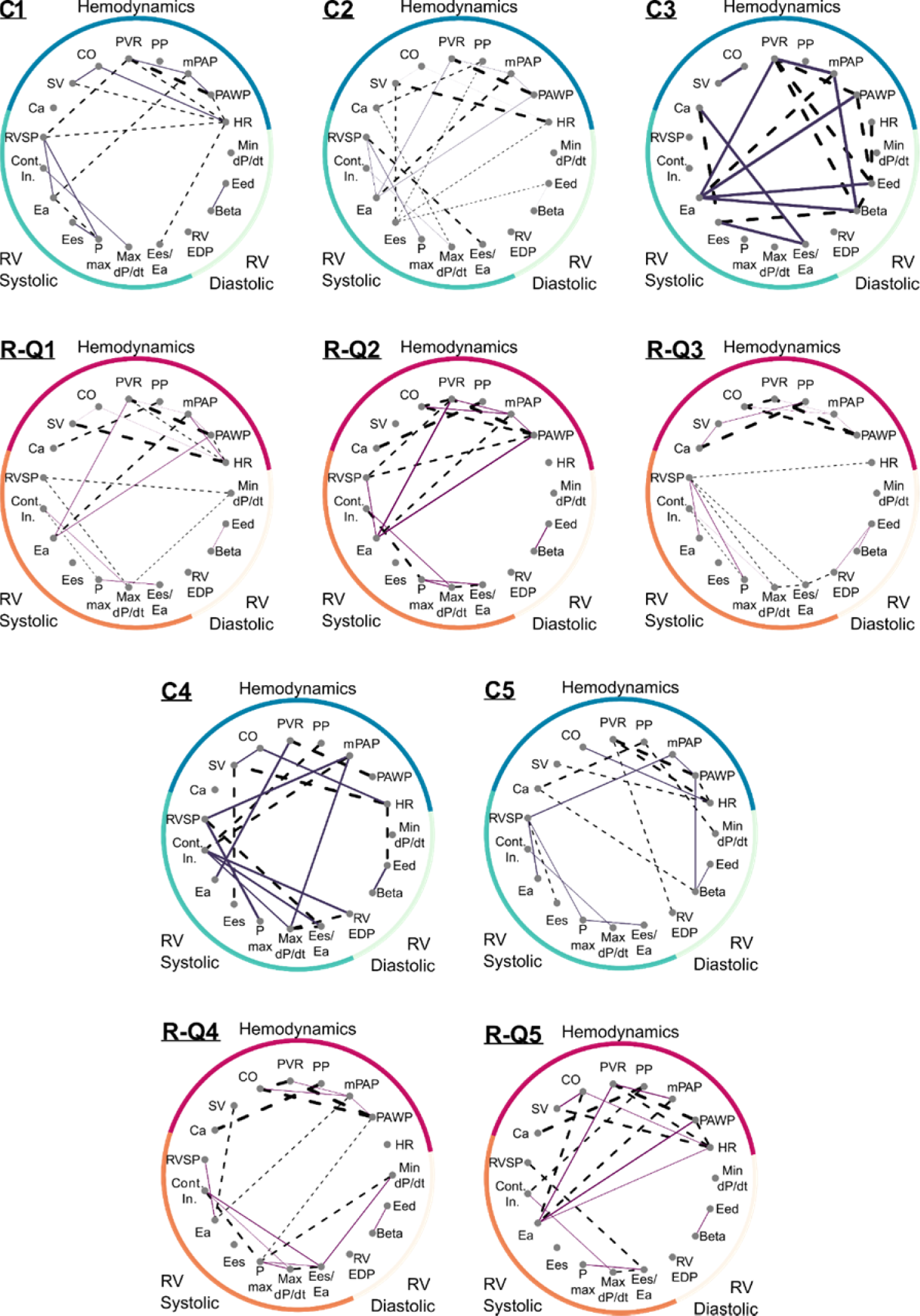
Network analysis to investigate variable-variable interactions in the Cluster and PVR quintile groups. Comparison of variable-variable interactions within the Cluster groups (C1 to C5) and PVR quintiles (R-Q1 to R-Q5). Edge pattern reflects whether the partial correlation is positive or negative (solid = positive, dashed = negative). Edge weight is proportional to the partial correlation values for each variable-variable pair. The interactions within Hemodynamics, RV systolic and RV diastolic variables change depending on the Cluster and PVR quintile group. In cluster 1 (C1), the variable-variable interactions are primarily within the Hemodynamics or within the RV systolic variables with little cross-talk. Cross-talk between variable groups increases in cluster 2 (C2) with a further increase in Cluster 3 (C3). The interactions in cluster 4 (C4) are more similar to cluster 2 where the interaction in cluster 5 (C5) are more similar to Cluster 1. The variable-variable interaction in the PVR quintiles follow more of an afterload-dependent change. In the first PVR quintile (R-Q1), the variable-variable interactions are primarily within the Hemodynamics or within the RV systolic variables. These interactions intensify in the second PVR quintile (R-Q2). The interaction pattern changes in R-Q3 with more isolated interactions within hemodynamic and RV systolic variables. The cross-talk between the RV systolic and RV diastolic variables increases in R-Q4. In R-Q5, the variable-variable interactions are primarily within the hemodynamics variables with cross talk to Ea.

Alternatively, variable-variable interactions in the PVR quintile groups are more reflective of an afterload-dependent continuum (**Figure 4**). In the first two PVR quintiles (R-Q1 and R-Q2, low afterload), interactions primarily occur within hemodynamic variables (PVR, mPAP and PAWP). While there are interactions observed between min dP/dt, RVSP and max dP/dt, the primary cross-talk is between Ea and pulmonary pressure. In R-Q3 (intermediate afterload), the cross-talk between variable types is lost. In both R-Q4 and R-Q5 (high afterload), there is increased cross-talk among variable types. Specifically, in R-Q4, the interactions between RV systolic and RV diastolic variables increases. In R-Q5, the variable-variable interaction between Ea and the hemodynamic variables reverts back to a pattern similar to that in R-Q2. In comparison to the clusters, variable-to-variable interactions in the PVR quintiles exhibit more of a continuum with increased afterload.

### Predictors of Mortality

There were 35 deaths during the follow-up period (median: 4.6 years, range 0.6 – 8.1 years). One and three-year survival was 90.5% and 67.4% respectively. As individual variables, Mean PA pressure, Pulse pressure, PA compliance, PVR, Ea and min dP/dt significantly associate with mortality after adjusting for age, sex and BMI (**Table 2**). Univariable Cox regression analysis showed that Ea (AUC: 0.681) and mPAP (AUC: 0.694) are better predictors of mortality compared to other hemodynamic and RV function variables (**Supplemental Figure 4**). In a multivariable model including mPAP, Ea, and adjusting for age, sex and BMI, mPAP was identified as an independent predictor of mortality. In the participants with PH (mPAP>25 mmHg), Ea (AUC: 0.664) and mPAP (AUC: 0.688) remained as variables that associated with mortality with the highest AUC (**Supplemental Table 6**). In the PH specific multivariable model (mPAP and Ea adjusting for age, sex and BMI), mPAP was again an independent predictor of mortality. Participants with an Ea > 0.70 mmHg/ml or a mPAP > 41 mmHg have increased mortality (**Figure 5A and B**).

**Figure 5:**
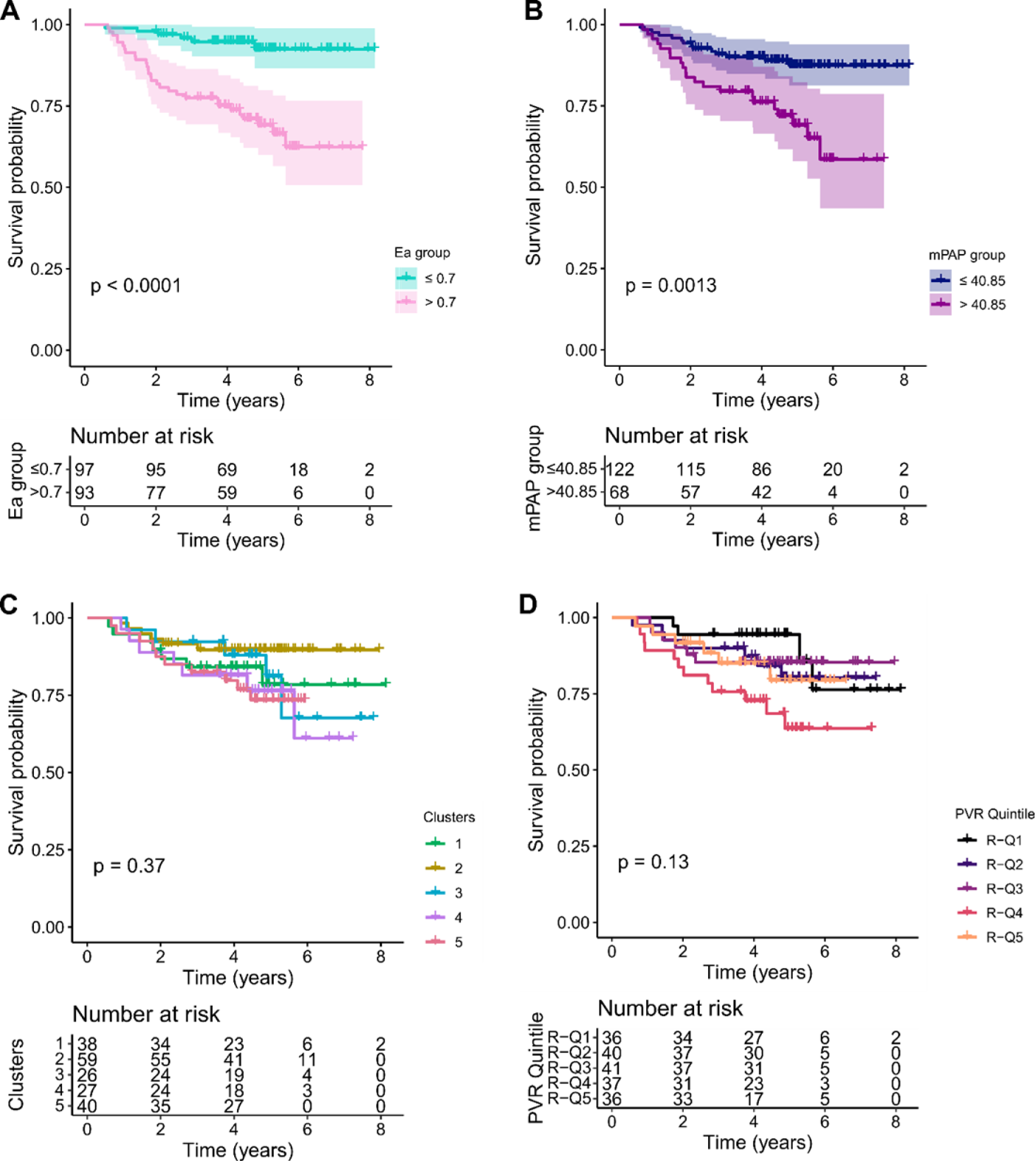
Association of Clusters and PVR quintiles with mortality. All-cause mortality survival analysis across RV function clusters **(A)** RV function variable Ea was significantly associated with survival over time. **(B)** Hemodynamic variable mPAP was significantly associated with survival over time. Kaplan Meier curves show no association with survival over time to follow-up for k-medoids derived RV function clusters (C1, C3, C4, C5 vs C2) and **(D)** PVR quintiles are significantly associated with survival over time (R-Q2, R-Q3, R-Q4, R-Q5 vs R-Q1). Log-rank test was used to determine significant differences across groups in Kaplan Meier.

**Table 2:** Cox proportional hazards ratios for survival. Univariate cox regression analysis and multivariate analysis when adjusted for age, sex, and BMI for significant variables across entire cohort showing the risk of death from the time of RHC to last date of follow-up. Multivariate models are composed of the significant univariate variables with the greatest AUC in ROC plots for hemodynamics or RV function. * denotes significant variables.

|  | Unadjusted Univariable<br>(p<0.1) |  | Adjusted for age+sex+BMI<br>Univariate (p<0.05) |  | Multivariable (p<0.05) |  |
| --- | --- | --- | --- | --- | --- | --- |
|  | 95% CI for HR | p-value | 95% CI for HR | p-value | 95% CI for HR | p-value |
| **Hemodynamics** |  |  |  |  |  |  |
| Mean PAP | 1.03 (1.01-1.05) | 0.002* | 1.04 (1.03-1.05) | 0.0004* | 1.05 (1.02-1.09) | 0.004* |
| Cardiac |  |  |  |  | -- | -- |
| Output | 0.98 (0.81-1.20) | 0.9 | 0.96 (0.77-1.19) | 0.69 |  |  |
| Pulse |  |  |  |  | -- | -- |
| Pressure | 1.03 (1.01-1.05) | 0.001* | 1.03 (1.02-1.04) | 0.001* |  |  |
| Ca | 0.69 (0.54-0.89) | 0.003* | 0.68 (0.54-0.81) | 0.003* | -- | -- |
| PVR | 1.083 (1.01-1.16) | 0.02* | 1.13 (1.09-1.17) | 0.003* | -- | -- |
| **RV Function** |  |  |  |  |  |  |
| Ea | 1.58 (1.02-2.45) | 0.04* | 1.78 (1.54-2.01) | 0.02* | 0.67 (0.27-1.7) | 0.4 |
| Max dP/dt | 1.001 (1.00-1.00) | 0.07* | 1.00 (1.00-1.00) | 0.05 | -- | -- |
| Contractile |  |  |  |  | -- | -- |
| Index | 0.96 (0.88-1.05) | 0.41 | 0.97 (0.88-1.06) | 0.5 |  |  |
| Ees | 1.22 (0.83-1.79) | 0.3 | 1.29 (0.87-1.92) | 0.2 | -- | -- |
| Ees/Ea | 0.99 (0.67-1.47) | 0.97 | 1.00 (0.68-1.48) | 1.0 | -- | -- |
| RVEDP | 1.03 (0.99-1.07) | 0.1 | 1.04 (0.99-1.08) | 0.09 | -- | -- |
|  |  |  | 0.998 (0.998- |  | -- | -- |
| Min dP/dt | 0.999 (0.998-1.00) | 0.03* | 0.999) | 0.0098* |  |  |
| Log(Beta) | 1.674 (0.76-3.69) | 0.2 | 1.69 (0.77-3.71) | 0.2 | -- | -- |
| Eed | 1.2 (0.68-2.12) | 0.5 | 1.11 (0.64-1.92) | 0.7 | -- | -- |

\* denotes significant variables

Pairwise log-rank testing showed no significant differences in prognosis between Clusters (**Figure 5C, p>0.05 all clusters**) or PVR quintiles (**Figure 5D, p>0.05 all quintiles**). As this is a retrospective analysis, treatment differences could present a significant bias survival analysis. On Cox regression analysis, cluster 4 (HR=4.24(3.70-4.78), p=0.008) had increased mortality compared to Cluster 2 (referent) when adjusted for age, sex, and BMI. Similarly, two PVR quintiles (R-Q4 (HR=3.83(3.17-4.49), p=0.04) and R-Q5 (HR=3.76(3.09-4.42), p=0.047)) had increased mortality compared to R-Q1 (referent) when adjusted for age, sex, and BMI.

## Discussion

We identified five distinct RV subphenotypes using unsupervised consensus clustering and hemodynamic measures of RV function including measures of RV-PA coupling (**Figure 3B**). The first two clusters are characterized as having low afterload and preserved (C1) or reduced RV systolic function (C2). Cluster 3 has decreased RV function with elevated pulmonary pressure, decreased contractility, decreased RV-PA coupling but with high CO. Clusters 4 and 5 describe moderate (C4) and severe (C5) decrease in RV function with high afterload, increased RV diastolic stiffness, and decreased RV-PA coupling in cluster 5.

An NIH-NHLBI identified area of investigation is in understanding RV function across the spectrum of PVD clinical phenotypes.^2^ One recommendation from the NHLBI led workshop was to apply systems biology and network medicine to endophenotype RV dysfunction in pulmonary vascular disease for the RV-specific treatment targets. Clustering methods have successfully been used to risk stratify patients with exercise intolerance^7^, identify immune phenotypes in PAH from proteomics^8^, and identify distinct phenotypes of IPAH^5^. However, clustering results are sensitive to collinearity between variables, randomness in the data, and the initial selection of the cluster centers. Components of pulmonary artery pressure are highly correlated^28^ and many hemodynamic measures of RV function are calculated values. Pearson correlation analysis was used to focus on important features of RV function while reducing co-linearity between variables.^7,13^ To account for inconsistencies and instabilities, we focused on consensus clustering methods that use validation statistics to identify clustering results that are reproducible^16,29^. In our dataset, K-medoids with a Pearson distance matrix outperformed hierarchical and k-means clustering to provide the most stable and consistent clusters over multiple runs. Using this rigorous clustering workflow, we identified five clusters that reflect varying degrees of RV dysfunction.

Clustering approaches with an RV-centric focus is one of the first steps in better endophenotyping the clinical presentation of RV dysfunction. RV dysfunction is a complex pathophysiology and non-linear process that is difficult to capture with individual parameters.^30^ Imaging parameters like RV ejection fraction^31^, end-systolic volume index^32^ and the ratio of stroke volume to end-systolic volume^20,33^ all significantly associate with mortality. Multi-beat conductance pressure-volume loop analysis is the gold standard for analyzing RV function and would be ideal for developing better RV endophenotypes.^34,35^ However, clustering approaches rely on large datasets and conductance pressure-volume analysis is limited to specialized centers and smaller cohorts. Clinical single-beat pressure volume loop analysis using RHC derived stroke volume, we are able to estimate RV contractility, arterial afterload and RV-PA coupling in a relatively large cohort (n=190 participants).^18,19,33^ By applying unbiased clustering approaches to the single-beat and hemodynamic measures of RV function, we identified 5 novel RV clusters that have varying degrees of afterload, contractility and RV-PA coupling.

When we took a more traditional approach and split the cohort based on increased afterload with the PVR quintiles, markers of RV dysfunction changed in an afterload dependent fashion (**Figure 3B, PVR quintiles**). Signs of dysfunction started to appear with a PVR > 3.2 WU (R-Q3) with altered end-diastolic elastance, PA compliance and increased RV contractility (Supplemental Table 5). In contrast to the Clusters, there were no significant differences in Ees/Ea across the PVR quintiles. These findings could be important with the new ESC/ERS definition of PH that includes a PVR cut-off of 2WU. The new PVR cut-off is based on the upper limit of normal for PVR and the lowest prognostically relevant threshold of PVR.^36–38^ However, it is important we quantify RV function in participants with a PVR between 2-3 WU in the development of RV-centric models.

RV dysfunction is not limited to specific WSPH classifications. Right ventricular function can be disproportionately altered in subgroups of pulmonary hypertension due to underlying biology^39^ Our data shows that WSPH groups were well distributed across the clusters and PVR quintiles (**Figure 2D and 2E**). The distribution is likely a result of WSPH classifications are based on etiology and clinical presentation rather than right ventricular function.^40,41^ Right ventricular afterload is typically higher in patients with WSPH group 1 but there is still a broad spectrum of RV dysfunction across WSPH groups. Risk assessment has primarily focused on WSPH group 1 but RV dysfunction is a significant contributor to clinical worsening. It is expected that a comprehensive classification would naturally capture aspects of different phenotypes because of all of these variables are physiologically interdependent. However, recent RV imaging studies have highlighted the need for risk stratification methods to include specific measures of RV function in multi-domain risk profiling of patients with pulmonary arterial hypertension. ^32,42,43^ The addition of RV function/imaging to current risk assessments help to discriminate intermediate and high-risk profiles. ^32,43^

Based on the analysis of variables, Cluster 3 presented several distinct and noteworthy features including reduced Ees/Ea despite a low PVR. The variable-variable network for Cluster 3 showed increased interconnectivity between systolic, diastolic and hemodynamic variables that was seen in other clusters or the PVR quintiles. Another unique feature of the Cluster 3 is high cardiac output (Table 1) that could be due to a number of factors including accompanying health conditions, prostacyclin therapy or obesity. The high cardiac output could be the results of portopulmonary hypertension, hyperthyroidism, anemia, chronic hypercapnia, liver disease, or congenital heart disease.^44^ Cluster 3 did have the highest percentage of participants with high PAWP (>15 mmHg) (7/15 participants; 46.7%). However, only 1 participant in the cluster (6.7%) was classified as WSPH group 2 suggesting other factors are contributing to the elevated wedge pressure. One effect of prostacyclin therapy is increased cardiac output and 5/15 participants in cluster 3 were on parenteral therapy. None of the participants in Cluster 3 had cirrhosis or portopulmonary hypertension and a high percentage (5/15) had connective tissue disease. Using BMI as a surrogate of obesity, cardiac output did not significantly associate with BMI (**Supplemental Figure 5**). A larger cohort is needed to identify if the increased cardiac output is a defining feature of C3 or just product of the cluster partitioning of the cohort.

### Clinical Relevance

Clustering participants allowed for the identification of novel RV phenotypes that consider systolic and diastolic RV function independent of WSPH classifications. The characteristics of RV function and the variable-variable interactions within the Clusters are different than those in the PVR quintiles. Additional work is needed to make better RV-centric risk stratification models as cluster 4 had an increased mortality risk (Table 3: HR: 4.24[3.70-4.78]) compared to cluster 2 but overall the Clusters did not associate with outcomes in a log rank test. Clinical application of these models is currently premature but these RV-centric clusters can help us better understand the hemodynamic presentation of RV dysfunction. The clustering variables that were used are all accessible from standard clinical tests without the need for specialized measurement equipment like conductance pressure-volume catheters^45^ or CMR imaging.

**Table 3:** Cox proportional hazards ratios for survival in clusters and PVR quintiles. Univariate cox regression analysis for clusters and PVR quintiles and cox regression analysis when adjusted for age, sex, and BMI. Cluster C2 was used as a referent for the clusters, and PVR quintile R-Q1 was used as a referent for the PVR quintiles. * denotes significant variables.

| Clusters | Unadjusted (p < 0.1) |  |  | Adjusted for age+sex+BMI (p<0.05) |  |
| --- | --- | --- | --- | --- | --- |
|  | PVR (WU) | 95% CI for HR | p-value | 95% CI for HR | p-value |
| C2 (referent) | 2.9 ± 1.5 | 1.0 | - | 1.0 | - |
| C1 | 2.9 ± 1.5 | 2.15 (1.59-2.70) | 0.17 | 2.37 (1.81-2.93) | 0.12 |
| C3 | 3.8 ± 1.1 | 2.23 (1.53-2.94) | 0.26 | 2.20 (1.47-2.92) | 0.28 |
| C4 | 7.9 ± 4.2 | 3.27 (2.75-3.80) | 0.025* | 4.24 (3.70-4.78) | 0.008* |
| C5 | 8.7 ± 3.5 | 2.64 (2.13-3.16) | 0.06 | 2.63 (2.10-3.16) | 0.07 |
| **PVR quintiles** | **PVR (WU)** |  |  |  |  |
| R-Q1 (referent) | 1.5 ± 0.4 | 1.0 | - | 1.0 | - |
| R-Q2 | 2.5 ± 0.3 | 0.58 (-0.39-1.49) | 0.55 | 0.54 (-0.38-1.45) | 0.5 |
| R-Q3 | 4.0 ± 0.5 | 2.35 (1.67-3.03) | 0.21 | 2.08 (1.40-2.76) | 0.3 |
| R-Q4 | 6.0 ± 1.0 | 4.27 (3.62-4.91) | 0.02* | 3.83 (3.17-4.49) | 0.042* |
| R-Q5 | 11.5 ± 3.7 | 3.38 (2.72-4.03) | 0.06 | 3.76 (3.09-4.42) | 0.047* |

\* denotes significant variables

Assessment of Pmax and Ees currently requires post processing of RV pressure waveforms that is not currently part of standard assessments.^46^ However clinical assessment of systolic, diastolic and RV-PA coupling from standard RV pressure waveforms allows for a broader application of the findings.^19^ In an exploration of decision tree models for predicting cluster assignment, the variables dP/dt, Ea, Eed and RVEDP were able to predict cluster assignment with 78% accuracy (**Supplemental Figure 3**). The search for the optimal collection of variables to inform therapeutic decisions is an ongoing endeavor to improve outcomes in patients with pulmonary hypertension.

### Limitations

The study includes a number of limitations The small size and single-institution cohort maybe have contributed to the lack of significance between cluster assignment and survival may be the result of a small single-institution cohort, treatment bias and/or survival bias.^47^ Follow-up time was assessed as time between catheterization and death/last follow-up. This did not account for the time between diagnosis and index catheterization that is variable in prevalent patients. As a retrospective study without follow-up, it was not possible to evaluate differences in therapeutic responses on RV function. Inclusion of therapeutic changes on the clustering variables could provide additional phenotype characterization information in the development of RV subphenotypes. Cohort size limited some of the sub-group analysis specifically, WSPH group 5 was excluded from subgroup analyses due to an n of 2. Cohort size also impacted the application of supervised models like decision trees due to limitations on the test/train datasets and increased overfitting of the models.^48^ We reran the consensus clustering algorithms 1000 times with bootstrapping on each algorithm to improve validation and consistency. These methods should be replicated on a larger validation cohort to investigate these unique aspects of the RV subphenotypes with particular focus on variable-variable interactions and unique cluster characteristics.

## Conclusion

This study demonstrates that the application of consensus clustering to hemodynamic measures of RV function may help identify better endophenotypes of RV function. Five distinct subphenotypes (clusters) were identified that differ substantially in contractility and RV-PA coupling (Ees/Ea) We detected a high-flow low-function phenotype that was absent when using other PH stratification criteria. Phenotype evaluation based on the RV function variables may help in identifying patient disease status and can provide future insights into RV endotypes and personalized treatment options that may better suit an individual’s needs.

## Data Availability

Data is available upon request.

## Acknowledgements

None.

